# Low-Frequency Deep Brain Stimulation Promotes Long-Term Orofacial Motor Recovery After Traumatic Brain Injury: A Case Study

**DOI:** 10.64898/2026.11.04.26366993

**Authors:** Lilly W. Tang, Erinn M. Grigsby, Jordyn E. Ting, Arianna Damiani, Jonathan C. Ho, Sirisha Nouduri, Sara Trant, Nicolo Macellari, Veronica Fossati, Kevin M. Franzese, Theodora Constantine, Donald J. Crammond, Kelly Zdrowak, Kaila L. Stipancic, Jorge A. Gonzalez-Martinez, Elvira Pirondini

**Author notes:** These authors contributed equally (Kaila L. Stipancic, Jorge A. Gonzalez-Martinez, Elvira Pirondini).

## Abstract

Diffuse axonal injury (DAI), a subtype of traumatic brain injury (TBI), can disrupt corticobulbar pathways, leading to chronic deficits in speech and swallowing. We present a first-in-human case study demonstrating the long-term therapeutic potential of low-frequency continuous deep brain stimulation (DBS) of the motor thalamus to restore corticobulbar function in a patient with severe DAI and persistent dysarthria and dysphagia. The patient underwent bilateral motor thalamus DBS implantation one-year post-injury for treatment of post-traumatic action tremor, and stimulation was delivered chronically at 55 Hz over three years of follow-up. We assessed orofacial motor outcomes using kinematic analysis of facial movements, videofluoroscopic swallowing studies, and speech tasks to dissociate the immediate assistive effects of stimulation (DBS ON versus OFF) from its long-term therapeutic effects (DBS OFF over time). Continuous DBS led to progressive recovery of facial symmetry, resolution of laryngeal penetration, and marked improvements in respiratory and phonatory control and speech intelligibility. While improvements in gross orofacial motor function persisted in the absence of stimulation, fine articulatory control remained dependent on ongoing stimulation or adjunct speech therapy for optimal performance. Stimulation-induced improvements in cervical posture may also have contributed to enhanced biomechanics of swallowing and voice production. These findings provide early evidence that low-frequency motor thalamus DBS may promote therapeutic benefits and extend the window of functional recovery in chronic TBI patients.

## Introduction

Diffuse axonal injury (DAI), a subtype of traumatic brain injury (TBI), frequently affects long-range projection fibers such as the corticospinal and corticobulbar tracts, leading to permanent impairments in speech and swallowing, namely dysarthria and dysphagia, respectively. These deficits can cause dire consequences for quality of life, contributing to depression, malnutrition, aspiration pneumonia, and even death.^1–3^ In the United States alone, an estimated 5.3 million people live with TBI-related disabilities. Conservative estimates suggest that annually over 500,000 patients experience chronic dysarthria while more than 1.3 million suffer from persistent dysphagia.^4–6^ Because speech and swallowing are essential to patients’ autonomy, communication, and wellbeing, preserving these functions is a critical priority of TBI rehabilitation. However, recovery typically plateaus within 6–12 months from injuries^7^, and standard interventions such as behavioral speech-language therapy and swallowing exercises, rely largely on compensatory strategies rather than restoring neuromuscular control, yielding limited gains in the chronic phase.^8^ Therefore, there exists an urgent clinical need for novel therapeutic approaches to treat chronic TBI survivors with persistent corticobulbar deficits affecting facial movement, speech, and swallowing.

We recently demonstrated that continuous low-frequency (50-80 Hz) deep brain stimulation (DBS) of the motor thalamus, more specifically the ventralis intermediate (VIM) and ventral oralis posterior (VOP) nuclei, enhances corticospinal and corticobulbar excitability by recruiting thalamocortical afferent projections to the motor cortex.^9–10^ In patients with TBI and chronic dysphagia and dysarthria, this increased excitability improved volitional control of facial and oropharyngeal muscles, swallowing efficiency, and speech intelligibility and quality. However, our previous study assessed only the immediate effects of stimulation: facilitatory effects emerged upon activation of thalamic stimulation and ceased promptly when stimulation was discontinued, consistent with an assistive effect^10^. Whether chronic stimulation could also produce long-term therapeutic effects that persist in the absence of stimulation remained unknown. Because continuous DBS has proven safe for the long-term treatment of movement disorders such as Parkinson’s disease and essential tremor^11^, we reasoned that continuous low-frequency motor thalamus stimulation could be delivered safely with minimal adverse effects over years in TBI patients and might support sustained motor recovery. Specifically, we hypothesized that chronic stimulation would produce such long- term therapeutic effects. If so, function with DBS OFF should improve over time, and the immediate assistive effect of stimulation (ON vs. OFF) should change accordingly.

Here, we had the opportunity to test this hypothesis on a case study: a patient sustaining moderate dysphagia and profound dysarthria following severe DAI who underwent a chronic bilateral DBS implant in the motor thalamus to treat post-traumatic action tremor. We assessed long-term therapeutic effects of low- frequency motor thalamus DBS on corticobulbar functions including volitional facial movement, swallowing coordination and efficiency, and speech production. We showed that continuous stimulation over the course of 3 years restored facial symmetry, resolved penetrations during swallowing, and ameliorated respiratory and phonatory control of speech. Our study provides first-in-human evidence that long-term low-frequency DBS may serve as a therapeutic tool for restoring swallowing and speech in patients with chronic oromotor deficits following TBI.

## Results

### Participant Characteristics

A male in his 40s (TBI01) sustained a severe TBI from an unhelmeted motor vehicle accident. Upon initial presentation, TBI01 scored a Glasgow Coma Scale (GCS) of 3T. MRI demonstrated bilateral DAI involving the corticobulbar and corticospinal tracts with scattered subarachnoid hemorrhage **(Figure 1A)**, prompting emergent external ventricular drain and bolt placement for intracranial pressure management. Associated traumatic neurologic injuries included T6-T9 vertebral body fractures, a C7 spinous process fracture, L2-L4 transverse process fractures without cord compromise. All spinal fractures were stable and managed nonoperatively with cervical-collar and thoracic-lumbar-sacral orthosis brace. The ventriculostomy and bolt monitor were removed on hospital day 7. Due to severity of injury, TBI01 sustained severe swallowing impairments that required a tracheostomy for respiratory support and a percutaneous endoscopic gastrostomy (PEG) tube placed at 2 weeks post-injury. Over a 1-month hospitalization period, TBI01 demonstrated mild gradual improvement to GCS 8 before discharging to acute inpatient rehabilitation. Over 4 months of inpatient rehabilitation, including up to 3 hours daily of physical therapy (PT), occupational therapy (OT), and speech therapy (ST), TBI01 was successfully weaned to a modified oral diet and deemed safe for discharge home with weekly outpatient rehabilitation follow-up. At discharge, he improved to a GCS of 9, though he continued to experience bilateral upper-limb paresis, severe facial paresis, moderate dysphagia, and profound dysarthria per SLP bedside evaluation.

**Figure 1.**
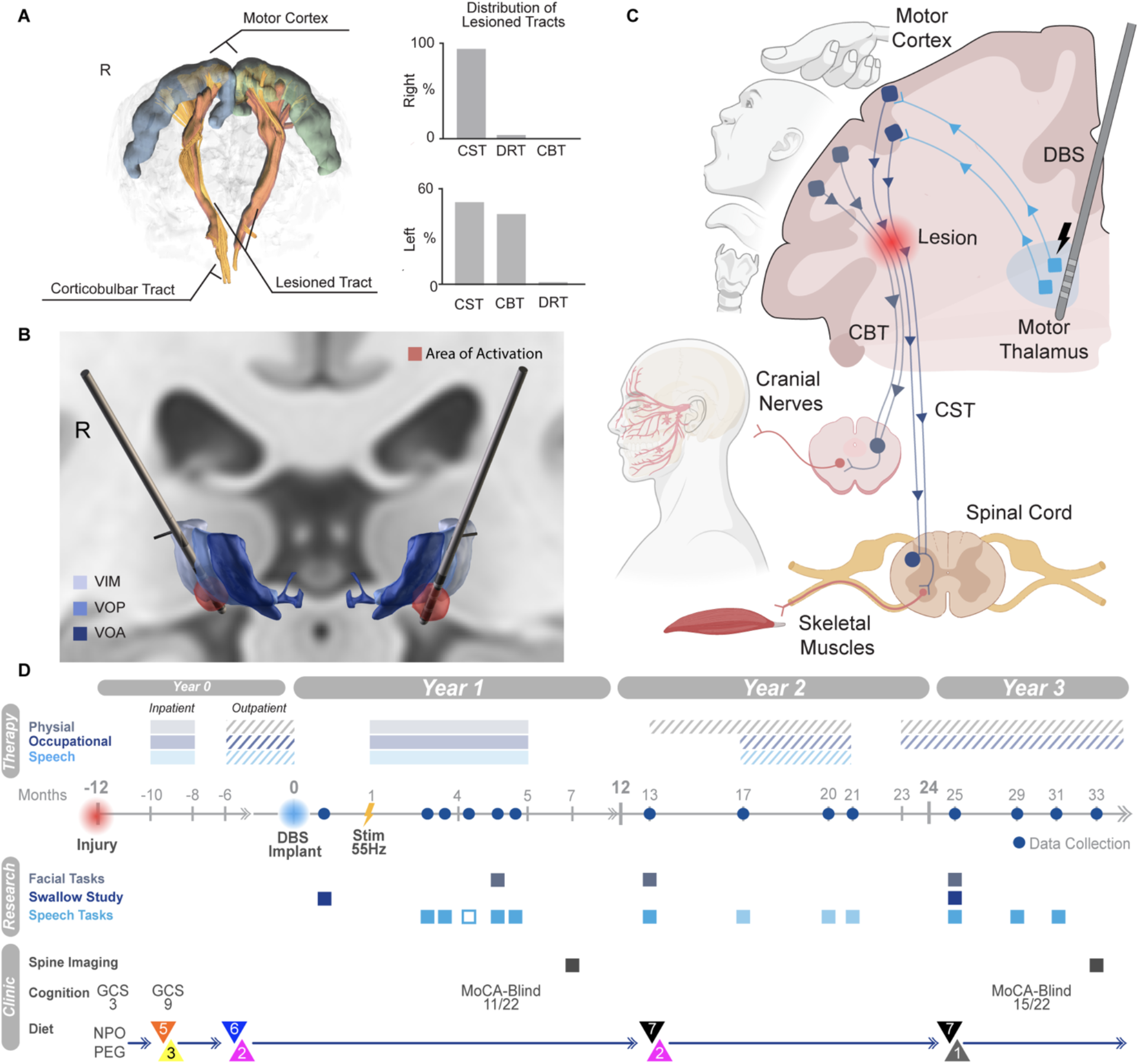
| Clinical design. **(a)** *Left*: Tractography with lesioned tracts highlighted in red. *Right:* Percentage distribution of lesioned tracts for right and left hemispheres. CST represents corticospinal tract, CBT for corticobulbar tract, and DRT for dentato-rubro-thalamic tract. **(b)** Implanted DBS lead localization relative to motor thalamus nuclei: ventralis intermedius (VIM, navy), ventralis oralis posterior (VOP, royal blue), and ventralis oralis anterior (VOA, sky blue). Area of stimulation activation denoted in red. **(c)** Schematic of mechanism of action. The motor thalamus (light gray) projects excitatory connections to the face/hand representation of the primary motor cortex (M1). Low-frequency electrical stimulation of the motor thalamus increases excitation of the face/hand motor cortex to cranial nerves and spinal cord, consequently potentiating muscles associated with the main facial articulators and upper limb skeletal muscles via projections through the CBT (dark gray) and the CST (dark blue), respectively. **(d)** Timeline of clinical study from initial injury to latest session of reported data. Years since DBS implant from 0 to 3 are depicted on top, with months from implant labelled on timeline. Therapy status is color coded with respect to physical, occupational, and speech therapy. Inpatient therapy, denoted in solid colors, entailed up to 3 hours per day, 6 days per week, whereas outpatient therapy, denoted in striped colors, consisted of 1 hour sessions, 2 days per week. All data collection sessions are indicated directly on the timeline (blue circle) with specification of facial, swallowing, and speech tests color coded under the Research label. For speech tasks, sessions with only sentence tasks are outlined, sessions with only phoneme analysis are notated in light blue, and sessions with all tasks are in solid blue. Evaluation of cognition included Glasgow Coma Scale (GCS) and Montreal Cognitive Assessment for Blind (MoCA-Blind). Diet status by recommendation of SLP are indicated as either NPO with PEG tube feeds or within levels in accordance with the International Dysphagia Diet Standardisation Initiative (IDDSI): *Top* for solid and *Bottom* for liquid diet status.^12^

## Clinical study design

Following his TBI, TBI01 developed a medically refractory tremor affecting the left upper extremity. One year after the injury, he underwent bilateral implantation of DBS electrodes (Medtronic SenSight leads connected to a Percept PC system) targeting the VIM/VOP nuclei of the motor thalamus (**Figure 1B-C**). We previously demonstrated that low-frequency (50-80 Hz) stimulation of the VIM/VOP nuclei induces immediate post-synaptic excitation of the cortico-spinal and cortico-bulbar motor neurons within the primary motor cortex^9^. This increased excitation facilitated immediate improvements of facial movement, swallowing, and speech in TBI01 along with other human subjects.^10^ Importantly, the stimulation facilitation was larger when stimulating at low-frequency (50-80 Hz) as compared to clinical standard frequency used to suppress tremor (130Hz). Therefore, therapeutic programming included both conventional high-frequency settings and a low-frequency range. After trialing both stimulation parameters, TBI01 chose low-frequency stimulation at 55 Hz (**Supplementary Table 1,** **Figure 1D)** and remained on this setting for the remaining 3 years. Stimulation was turned OFF only for acute testing of immediate effects and overnight for battery conservation while asleep. This provided the opportunity to characterize the sustained clinical and physiological effects of low-frequency motor thalamus stimulation over the three years following implantation. At 55 Hz, TBI01 achieved clinically meaningful suppression of his left upper extremity tremor, comparable to that obtained with conventional high-frequency (130 Hz) settings, while additionally conferring the facial, swallowing, and speech benefits previously described.^10^ Stimulation at this frequency was well tolerated throughout the 3-year follow-up period, with no serious adverse events reported. The only adverse effect noted was transient facial paresthesia lasting 2–5 minutes following initial stimulation parameter adjustments, which resolved spontaneously without intervention and did not recur with stable, chronic stimulation.

To monitor the effect of stimulation over time, we analyzed facial and oropharyngeal motor performance data collected during structured behavioral experiments along with patient and caregiver reports collected over the 3 years from DBS implantation (**Supplementary Table 2**). Specifically, we evaluated: 1) volitional facial control using standardized facial motor tasks, 2) swallowing using videofluoroscopic swallow studies, and 3) speech motor control during speech tasks. For each task, we assessed i) immediate effects of stimulation by comparing behavioral performances with DBS ON versus OFF within the same session, and ii) ong-term effects of stimulation by comparing performances over time with DBS OFF.

## Motor thalamus DBS restores facial symmetry

TBI01’s left-hemispheric CBT lesion (>45% of tracts lesioned, **Figure 1A**) caused an orofacial muscle weakness resulting in a severe right-sided facial paresis and a marked facial asymmetry. To assess the effect of motor thalamus DBS on facial paresis and asymmetry, we evaluated movement performances over time while TBI01 performed standardized facial motor tasks: moving between instructed (tongue in/out, smile, mouth open/close, and lip pucker) and neutral facial expressions as large and as fast as possible, with and without stimulation. Videos of TBI01 performing the tasks were recorded for kinematic analysis. Upon broad visual inspection, we observed a clear increase in orofacial symmetry over time (**Figure 2A- B**). We first quantified the right mouth aperture at maximum expression of each facial task measured from three facial articulators (right corner of mouth, upper lip, and lower lip). As expected, low-frequency motor thalamus stimulation produced immediate significant increase in right-sided mouth aperture compared to no stimulation for three out of four tasks (Tongue InOut, Smile, and Mouth OpenClose). Moreover, DBS also promoted a significant increase in right-sided mouth aperture over time across both stimulation ON and OFF conditions for these same tasks (**Figure 2C**). Given this marked improvement in right-sided facial paresis, we then sought to assess immediate and long-term effects of low-frequency stimulation on facial symmetry by computing the ratio of right to left mouth aperture (**Figure 2D**). As we previously reported,^10^ in year 1 DBS significantly reduced the severe orofacial asymmetry across all four facial tasks (Tongue InOut 0.56 to 0.82; Smile 0.58 to 0.77; Mouth OpenClose 0.56 to 0.93; Lip Pucker 0.59 to 0.72). By year 2, stimulation-related improvements were evident only in two tasks in which asymmetry persisted (Tongue InOut from 0.78 to 1.0; Mouth OpenClose 0.89 to 1.0). By year 3, the aperture ratio approached 1 for all tasks, even with stimulation OFF (Tongue InOut 0.99; Smile 1.0; Mouth OpenClose 0.97; Lip Pucker 0.83), suggesting that the severe orofacial asymmetry was restored (**Figure 2E**). Unsurprisingly, because of this restoration, there were no longer evident immediate effects of the stimulation by year 3.

**Figure 2.**
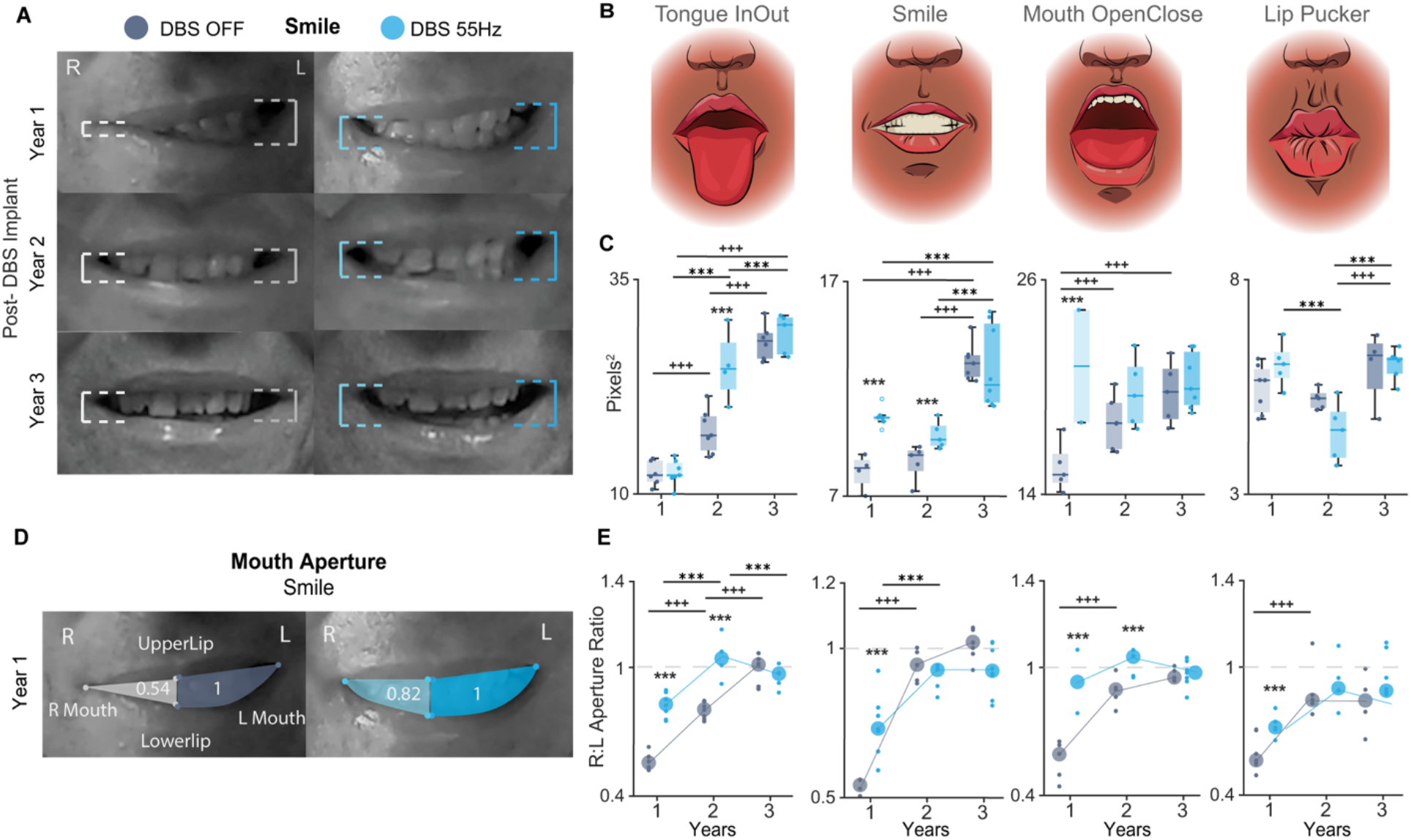
| Long-term motor thalamus stimulation restores orofacial symmetry. **(a)** Frame captures from video showing TBI01 maximum range of motion of the lips during smile task for DBS OFF (gray) and DBS ON (blue) conditions over the 3 years. Right side is colored in a lighter shade than the left. **(b)** Schematic of facial tasks Tongue InOut, Smile, Mouth OpenClose, and Lip Pucker. **(c)** Measurement of maximum area of active mouth aperture across facial tasks over the 3 years for DBS OFF (gray) and DBS ON (blue). **(d)** Example area of visualization calculated from 3 representative kinematic markers: upper lip, lower lip, and corner of mouth between right and left sides during smile task for DBS OFF (gray) and DBS ON (blue) conditions. Left-to-Right ratio depicted relative to L-side as 1. **(e)** Ratio of mouth aperture of right side to left side for the 3 years since DBS implant for facial tasks tongue in/out, smile, mouth open/close, or lip pucker facial expressions. Each small dot corresponds to one trial, with the large dot representing the average over trials. DBS OFF is indicated in gray and DBS ON in blue. Statistical significance was assessed with two-tail bootstrapping with Bonferroni correction: significance between DBS OFF and ON within the same sessions are indicated with p<0.05 (*), p<0.01 (**), p<0.001(***), significance across years for DBS OFF with p<0.05 (^+^), p<0.01 (^++^), p<0.001(^+++^), and significance across years for DBS ON with p<0.05 (*), p<0.01 (**), p<0.001(***).

Overall, these results suggest that low-frequency motor thalamus DBS over time strengthens muscle activation, resolving facial paralysis that persisted even when stopping the stimulation.

### Motor thalamus DBS improves oropharyngeal swallowing coordination and efficiency over time

To assess the effects of motor thalamus DBS on TBI01’s swallowing impairments, we conducted videofluoroscopic swallow study (modified barium swallow test)^13^ at year 1 without stimulation and at year 3 with DBS ON and OFF. We also tracked speech-language-pathologist (SLP) diet recommendations and patient reports over the same period. Two SLPs who were blinded to the experimental design and stimulation conditions, independently scored each videofluoroscopic study using the Penetration Aspiration Scale (PAS)^14^ and the Dynamic Imaging Grade of Swallowing Toxicity (DIGEST).^15^ A PAS score >2 (8- point scale) indicates unsafe swallowing. The DIGEST score is a 5-point scale that assigns a global rating of pharyngeal swallowing function according to the interaction of the safety and efficiency profile (0=no pharyngeal dysphagia, 1=mild dysphagia, 2=moderate dysphagia, 3=severe dysphagia, 4=life threatening dysphagia). Because TBI01 was instructed to clear each bolus in a single swallow whenever possible, any amount of bolus remaining after the initial swallow was graded as an efficiency deficit. To further characterize oropharyngeal coordination, the SLPs graded coordination of each oropharyngneal structure across swallowing phases. From this evaluation, we derived a 4-level ordinal scale across 12 components: mastication, bolus cohesion, anterior-posterior transfer, oral residue, velarpharyngeal closure, tongue retraction, epiglottic inversion, hyo-laryngeal excursion, laryngeal vestibule closure, pharyngeal stripping, pharyngeoesophageal segment distention and duration (0 = complete, 1 = adequate, 2 = reduced, 3 = incomplete, total score of 36) (**Figure 3C**).

**Figure 3.**
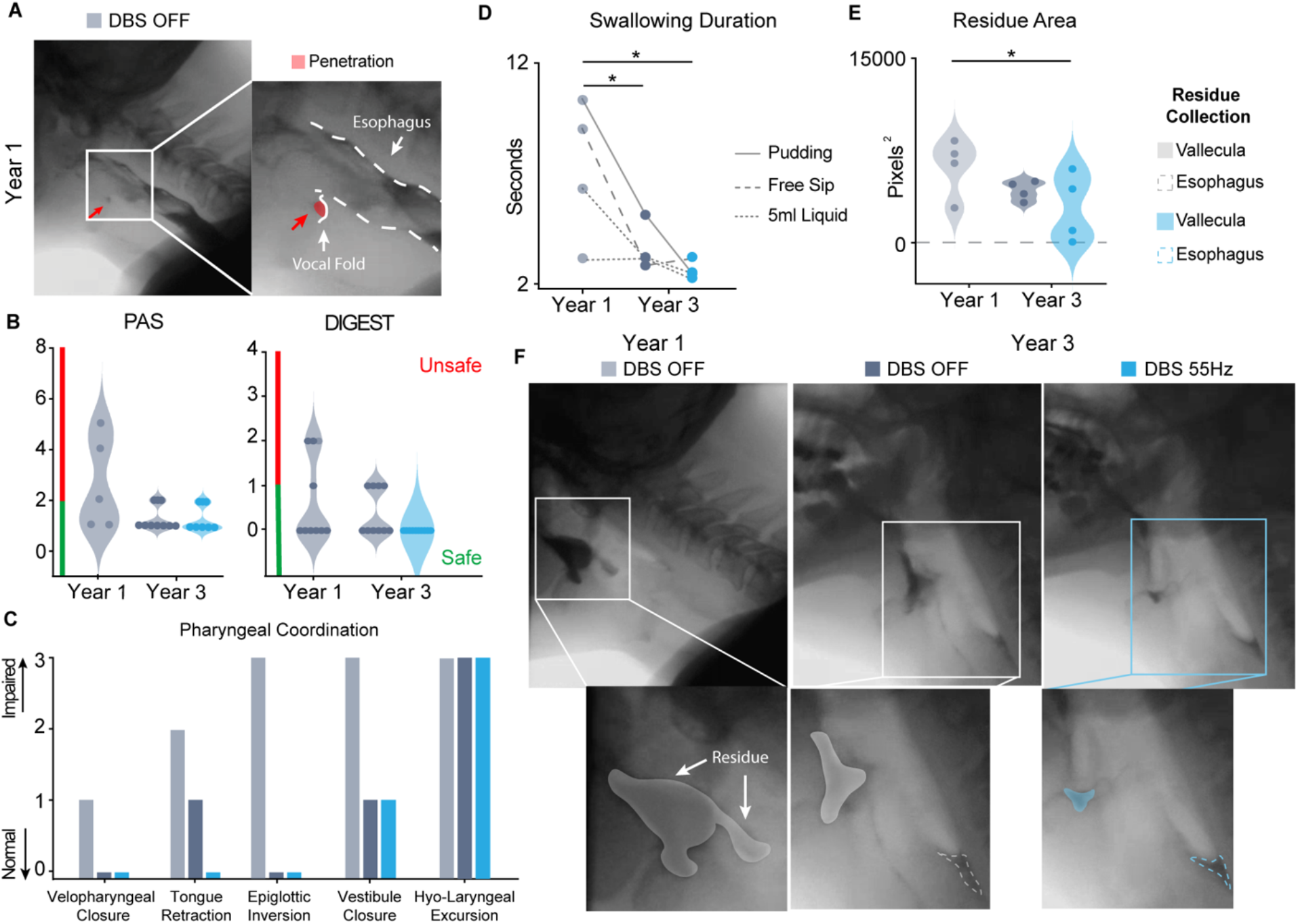
| Longitudinal assessment of motor thalamus stimulation reveals clinically-significant improvement in swallowing function. **(a)** Area of penetration from frame captures of modified barium swallow test conducted 1 year post-injury and DBS OFF. Area of deep penetration to vocal fold is highlighted by the red arrow and shaded in red. Please note that by year 3 there was no penetration. **(b)** *Left:* PAS of all trials of the modified barium swallow test at year 1 and year 3 post-DBS implant. *Right:* DIGEST at year 1 and year 3 post-DBS implant. Both scores are rated by blinded SLPs (n=2, **Supplementary Table 3**). **(c)** Graded score of modified barium swallow report for the pharyngeal phase of swallow coordination across year 1 and year 3 with DBS ON and OFF. **(d)** Total duration of matched trials of videofluoroscopic swallow study across years and stimulation conditions. Note two repetitions were performed for 5mL liquid trials. **(e)** Maximum area of vallecula and esophagus bolus remnants quantified from frame captures of phase-matched fluoroscopic capture of videofluoroscopic swallow study trials between year 1 and year 3 for conditions DBS OFF and ON. **(f)** *Top:* Frame captures of phase-matched fluoroscopic capture of videofluoroscopic swallow study for a representative trial of maximal bolus remnant within phase-matched pudding trial at year 1 and year 3 for DBS OFF (gray) and ON (blue). Positional difference in framing matched to size in reference to vertebrae anatomical landmarks. *Bottom:* Boluses remnants are shaded or outlined for reference. For all panels, DBS OFF at year 1 is labeled in light gray, DBS OFF at year 3 in dark gray, and DBS ON at year 3 in blue. Statistical significance for panels (d-e) was determined through matched one-way ANOVA, p<0.05 (*).

At year 1, TBI01 presented with moderate oral and pharyngeal dysphagia characterized primarily by reduced hyo-laryngeal excursion and incomplete laryngeal vestibule closure. Critically, he exhibited deep laryngeal penetration, with material contacting the vocal folds without being ejected, that was not mitigated by compensatory strategies (**Figure 3A**). These deficits were captured by a PAS score of 5/8 and a DIGEST score of 2/4, suggestive of moderate dysphagia (**Video 1,** **Figure 3B**). Accordingly, he remained on a modified diet (IDDSI level 6 soft/bite size; level 2 midly thick liquids). By year 3, swallowing function had improved substantially even in the absence of stimulation. Although hyolaryngeal excursion remained reduced, velopharyngeal closure, epiglottic inversion, and laryngeal vestibule closure, all critical for airway protection, improved (**Figure 3C**, total score: 27/36 for year 1 and 17/36 for year 3, respectively). Consequently, deep penetration resolved, and PAS improved to 2/8, within the safe range. DIGEST improved to 1/4, indicative of mild dysphagia (**Video 1**, **Figure 3B, Supplementary Table 3**). These gains were reflected in TBI01’s diet, which was advanced to IDDSI level 7 (easy to chew) with level 1 (slightly thick) liquids. Importantly, at year 3, DBS continued to confer additional benefit. Specifically, while the PAS and DIGEST scores in year 3 all spanned the safe range of swallowing, TBI01 more consistently scored lower with DBS ON (1/8 and 1/4, respectively, **Supplementary Table 3**), indicative of persistent effect of stimulation on improving swallowing efficiency. Indeed, complete restoration of tongue retraction was only observed in the DBS ON condition. Importantly, across all VFFS, PAS and DIGEST scores were highly reliable between SLPs, with a weighted Cohen’s kappa of k = 0.737 for PAS and k = 0.615 for DIGEST, both suggestive of substantial interrater agreement.

We further assess the changes on swallowing efficiency by quantifying the duration of swallowing and the maximum amount of bolus spillage after the first swallow in the videofluoroscopic swallowing study.^10^ Dysphagia commonly results in delayed bolus transit and increased pharyngeal residue due to impaired strength and coordination of oropharyngeal musculature.^16^ Consistent with the clinical ratings, swallow duration was significantly shorter at year 3 than at year 1 (p=0.027, n = 4 tasks), indicating a faster, more efficient swallow (**Figure 3D**). Bolus remaining in the pharynx after the first swallow also decreased by 44% from year 1 to year 3 with DBS OFF (average area of 7800.6 pixels^2^ at year 1 compared to 4362.8 pixels^2^ at year 3, **Figure 3E–F**). Stimulation continued to confer additional benefit at year 3. Indeed, it reduced pharyngeal bolus remnants (p = 0.037, n = 4 tasks; **Figure 3E–F**), consistent with our previous report^10^.

Together, these results show that swallowing safety and efficiency improved substantially between year 1 and year 3 even without stimulation, while DBS ON provided an additional, immediate benefit on bolus clearance. Functionally, TBI01 progressed from a modified to a near-regular diet. Such late recovery of swallowing coordination, years after injury, is rarely reported in the TBI literature.^17,18^

## Chronic motor thalamus DBS enhances speech motor function

Because TBI01 had a tracheostomy at the time of discharge to acute rehabilitation, formal SLP evaluation of his speech was not possible until approximately 1 year post-injury, after decannulation and discharge from acute rehabilitation, when he presented for DBS evaluation. At that time, TBI01 presented with a profound motor speech impairment characterized by low vocal intensity, reduced respiratory support, and imprecise articulation with marked consonants distortions per SLP evaluation. To assess progress across each speech subsystem over time, we recorded speech testing sessions in which TBI01 repeated short 5 to 8-words phoneme-balanced sentences selected from the Assessment of Intelligibility of Dysarthric Speech (AIDS) database and mono- to triple-syllable words that included the full spectrum of American English phonemes and required different levels of coordination and placement of lips, soft palate, and tongue (**Supplementary Table 2**).

TBI01’s vocal intensity increased significantly and consistently over the first 2 years, both with DBS OFF and ON (**Figure 4A, Supplementary Figure 1A**). At year 1, his mean vocal intensity was 40.64±0.79 dB, well below normative values for age- and sex-matched healthy speakers (59.5(±3.7) dB^19^). With DBS OFF, vocal intensity plateaued by year 2 (mean 53.06 ± 1.33 dB; maximum 54.46 dB), whereas with DBS ON it continued to increase in year 3 (mean 53.81 ± 1.79 dB; maximum 55.85 dB). Thus, the immediate effect of stimulation, which was large at the beginning of year 1, diminished over time as OFF performance improved, but remained significant in some sessions of year 3. Similarly, the number of voice breaks within sentences decreased significantly from year 1 to year 3 in both conditions, with no significant difference between DBS ON and OFF within each year (**Figure 4B, Video 2**). Together, these results indicate that improvements in respiratory and phonatory control were driven primarily by long-term therapeutic effects of stimulation rather than by its immediate assistive effects.

**Figure 4.**
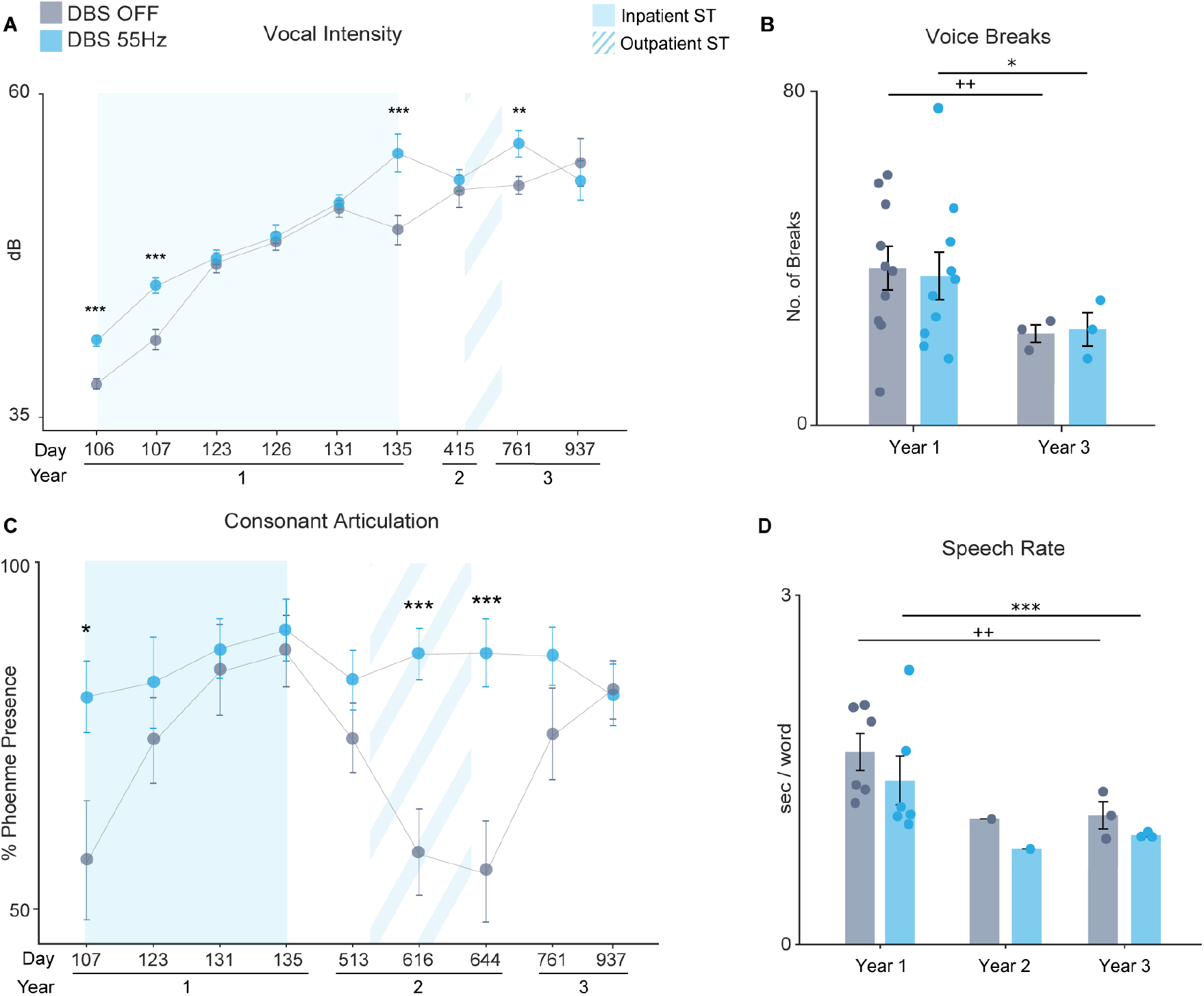
| DBS enhances respiration, phonation, articulation, and speech efficiency over time. **(a)** Speech intensity for DBS OFF and ON at 55 Hz conditions for single words tested across sessions over time. The top x-axis indicates days and bottom x-axis indicates year since DBS-implant. Light blue box indicates time when TBI01 received concurrent inpatient speech therapy. Light blue dashed box corresponds to the time when TBI01 underwent concurrent outpatient speech therapy. Note only sessions with same acquisition set-up were included for consistent matched comparison. **(b)** Bar plot of number of voice breaks detected in the sentence tasks for stimulation OFF and ON at 55Hz across year 1 and 3 post DBS-implant. Each dot corresponds to the number of voice breaks for one sentence tested. **(c)** Percentage of perceptual presence for consonants tested when DBS was ON at 55 Hz as compared to DBS OFF in single words tested across sessions over time. The top x-axis indicates days and bottom x-axis indicates year since DBS-implant. Note only sessions with consistent number of words tested (12-14 words) were included in matched analysis. **(d)** Bar plot of speech rate across three years for DBS ON and OFF conditions. Each dot corresponds to a single testing session. For all panels, DBS OFF condition is labeled in gray and DBS ON in light blue. Statistical significance was assessed with two-tail bootstrapping with Bonferroni correction: significance between DBS OFF and ON within the same sessions are indicated with p<0.05 (*), p<0.01 (**), p<0.001(***), significance across years for DBS OFF with p<0.05 (^+^), p<0.01 (^++^), p<0.001(^+++^), and significance across years for DBS ON with p<0.05 (*), p<0.01 (**), p<0.001(***). No statistical significance was calculated for Year 2 because we performed only one session of recordings.

In contrast to phonation, articulation with DBS OFF did not improve steadily over time but fluctuated across sessions (**Figure 4C, Supplementary Figure 1B**). These fluctuations tracked speech therapy status: OFF performance was higher during periods of concurrent speech therapy and showed marked variability in year 2, coinciding with TBI01’s transition from inpatient to outpatient speech therapy, when therapy intensity and consistency were reduced. In contrast, with DBS ON, the proportion of correctly produced consonants remained high and stable across years and was significantly greater than with DBS OFF, independent of speech therapy status (**Figure 4C**). These results suggest that fine articulatory control continued to rely on the immediate assistive effects of stimulation, or on concurrent behavioral therapy, to maintain optimal performance.

Finally, we assessed whether stimulation would improve speech rate, measured as the time required for TBI01 to complete speech tasks normalized by total number of words tested. We observed a significant decrease in utterance time per word between year 1 and year 3 (**Figure 4D**). Additionally, while not statistically significant, utterance time was consistently shorter in the DBS ON condition compared to DBS OFF within each year, suggesting a trend for DBS to increase speech rate towards the speed of natural conversation.^20^

Overall, these results show a dissociation between speech subsystems. Respiratory and phonatory control improved progressively and persisted in the absence of stimulation, consistent with a long-term therapeutic effect. In contrast, fine articulatory control remained dependent on ongoing stimulation or concurrent speech therapy.

## Improved cervical posture potential contribution to observed functional improvements

The pharyngeal phase of the swallowing and vocal phonatory control are supported by the activation of the neck and paraspinal muscles.^21,22^ At year 1, TBI01 exhibited forward head posture (FHP), a pathological posture indicative of neck muscle spasticity. Notably, cervical neck control has been recognized as an early predictor of recovery following severe TBI and is not generally observed to improve naturally in the chronic phase.^23^ Meanwhile, spasticity is well documented in literature as a chronic deficit that progressively worsens overtime, often requiring escalating intervention rather than resolving spontaneously.^24^ Thalamic stimulation has been previously reported to improve post-traumatic dystonia of cervical and upper-limb distributions.^25^ Similarly, we noticed a gradual change in TBI01 cervical posture over time that was not immediately apparent between stimulation ON versus OFF conditions. These observations prompted us to investigate the effect of chronic motor thalamus DBS on TBI01’s cervical postural.

To quantify postural changes, we first identified chin and chest kinematic markers from video recordings of TBI01 performing facial tasks across the three years. Because posture did not differ noticeably between stimulation conditions within sessions, we pooled DBS ON and OFF recordings. We then calculated the distance between the chin and chest markers as a measure of neck extension (**Figure 5A**). Notably, TBI01 showed a significant increase in chin-to-chest distance over the course of the 3 years (**Figure 5B**), suggesting an improvement in neck extension and cervical posturing.

**Figure 5.**
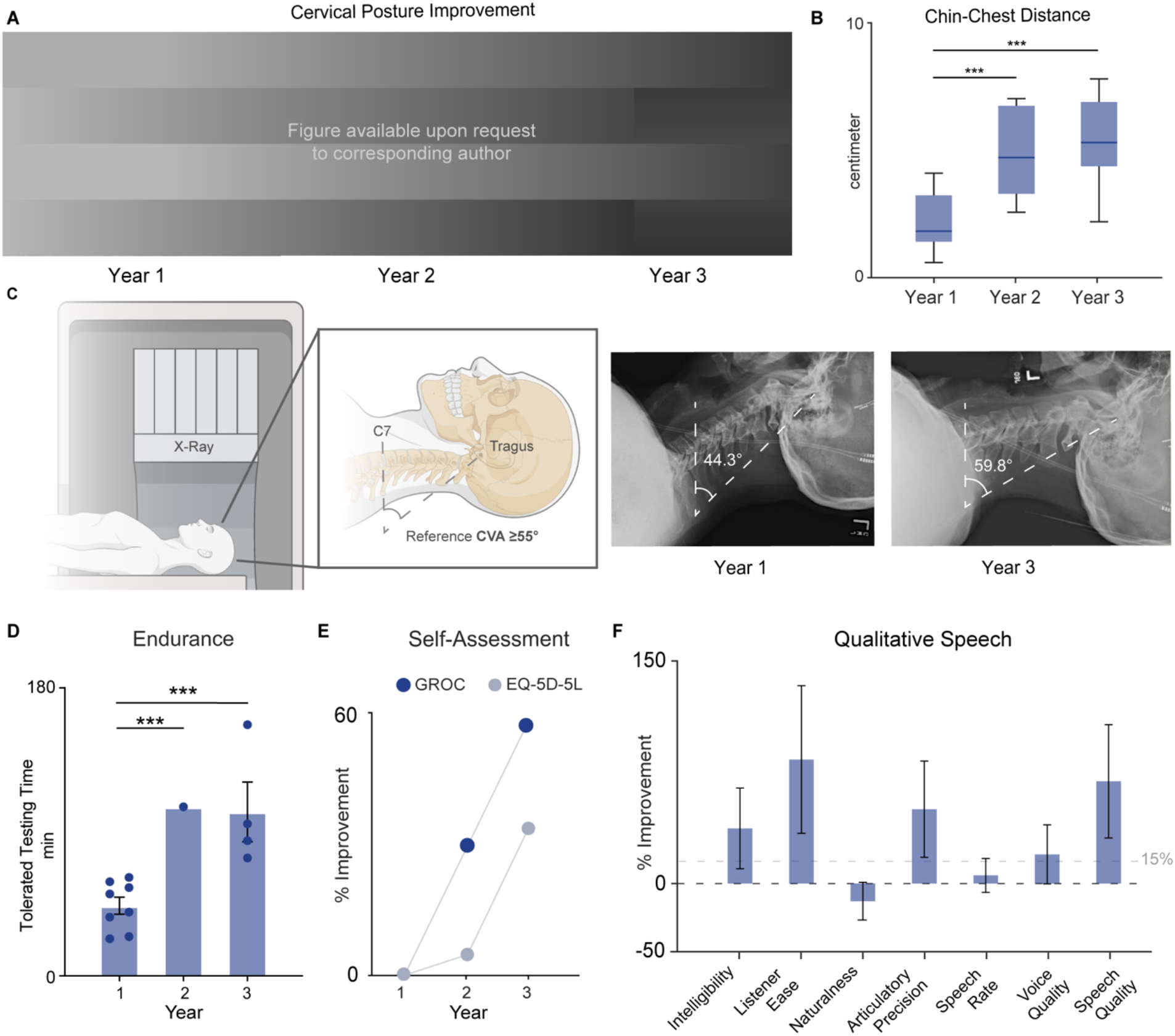
| Long-term motor thalamus DBS improves cervical posture and qualitative metrics. **(a)** Frame capture with labeled kinematic marker of chin and chest across three testing sessions across 3 years. Dotted line demonstrates chin-to-chest distance. **(b)** Boxplot of chin-to-chest distance for each year. For all panels, DBS OFF condition is indicated in gray whereas DBS ON in blue. **(c)** *Left:* Supine X-ray acquisition schematic for measurement of craniovertebral angle (CVA) calculated from horizontal level of the C7 spinal process to tragus of ear. *Right:* X-ray imaging of the cervical spine at year 1 and year 3 acquired with standard supine imaging protocol, with the back of the head in contact with the imaging table as much as possible. Note for year 1, TBI01 had significant difficulty maintaining contact with imaging table due to cervical spasticity that is resolved by year 3. **(d)** Endurance quantified by total tolerated testing time in sessions across three years, accounting for varying stimulation conditions. **(e)** Percentage of improvement in self-reported assessments of global rating of change (GROC) and EQ-5D-5L across three years post-TBI. **(f)** Percentage of improvement in qualitative speech metrics across sentence between year 1 and year 3 with no stimulation rated by naïve native English speakers (n=6). Dots represent values for individual rater across each sentence sample evaluated. Error bars indicate SEM. Grey dotted line demarcates minimal clinically important difference (>15%). Statistical significance was assessed with two- tail bootstrapping with Bonferroni correction: significance between across years are indicated with p<0.05 (*), p<0.01 (**), p<0.001(***).

We also measured the craniovertebral angle (CVA)^26^ from supine radiographic imaging acquired at year 1 and year 3 using the same imaging protocol. CVA is a standard metric of head and neck posture, derived from the angle between a horizontal line through the C7 vertebral level and a line from the C7 spinous process to the tragus of the ear. CVA ≥ 55° is considered healthy physiology whereas CVA ≤ 45° is considered severe FHP (**Figure 5C**).^27,28^ Imaging protocol was acquired with TBI01 supine and the back of his head in contact with the imaging table. At year 1, TBI01 had a CVA angle of 44.3°, indicative of severe FHP. Notably, TBI01 could not extend his neck to bring the back of his head completely in level at year 1 due to severe cervical spasticity. However, by year 3, the CVA angle was restored to a healthy range of 59.8°, significantly improving neck extension (**Supplementary Figure 2D**).

Forward head posture and diminished CVA has been mechanistically linked to worsening swallowing and voice production biomechanics.^29,30–32^ Given that this degree of cervical correction is not expected from natural recovery history alone and prior evidence of thalamic DBS improving post-traumatic cervical dystonia, ^23–25^ we propose that TBI01’s postural change reflect a stimulation-related effect that may have contributed to the observed improvements across speech intensity, phonation, and swallowing function.

## Motor thalamus DBS improves overall physical endurance, quality of life, and speech quality

To assess the broader impact of long-term DBS on TBI01’s overall quality of life, we first investigated the effect of stimulation on overall physical endurance in facial and oropharyngeal motor functions. For this, we recorded the total amount of testing time for each session. For all sessions, TBI01 performed behavioral assessments that required facial and oropharyngeal motor strength and coordination until his maximum tolerance was reached, as indicated by his request to stop testing. Therefore, an increase in tolerated time suggested an overall improvement in motor endurance. Interestingly, TBI01 exhibited a steady increase in tolerated testing time over the three years, corroborating the positive effect of long-term thalamic stimulation on physical endurance (**Figure 5D**).

Next, we collected self-reported surveys of global rating of change (GROC)^33^ and EuroQol EQ-5D-5L^34^ across three years post-TBI to assess the therapeutic impact of stimulation on health-related quality of life. TBI01 reported a consistent improvement on GROC increasing from +1 (a tiny bit better) to +3 (somewhat better) to +5 (quite a bit better), for year 1, 2, and 3, respectively. Similarly, EQ-5D-5L also improved over time, decreasing from 23/25 at year 1 to 15/25 at year 3, with more drastic improvement between year 2 to 3 (**Figure 5E, Supplementary Table 4**).

Finally, we evaluated the overall change in TBI01’s perceptual speech quality. For this, we recorded TBI01 speaking phoneme-balanced sentences at Year 1 and Year 3 and asked naïve blinded native-English speakers (n=6) to rate TBI01’s speech performance across various perceptual speech measures on a scale of 0-100 (0 as severe impairement and 100 as no deficit).^35,36,37^ Listeners noted an clinically significant improvement comparing year 3 to year 1 in intelligibility (31.6%), listener ease (71.1%), voice quality (16.7%), articulatory precision (42.6%), and overall quality of speech (58.6%) (**Video 2,** **Figure 5F**).^38,39^

## Discussion

This first-in-human longitudinal case study demonstrates that continuous low-frequency stimulation of the motor thalamus may drive clinically meaningful and durable recovery of orofacial motor function in the chronic phase after severe traumatic brain injury with diffuse axonal injury. Improvements were consistently observed across multiple cortico-bulbar domains, including facial symmetry, speech, swallowing, and cervical posture.

To our knowledge, this is the first report to document progressive improvements across orofacial and cervical motor functions beyond one-year post-injury in TBI. In typical recovery trajectory for severe TBI with DAI, improvements in swallowing and speech occur predominantly within the first 3-6 months post- injury, with deficits persisting beyond 12 months even with intensive rehabilitation.^4,17,40,41^ In our study, instead, continuous motor thalamus DBS successfully eliminated deep penetration to the level of the vocal folds and restored safe swallowing, in a patient with persistent moderate dysphagia 1 year post-injury in absence of active therapy. The resolution of deep penetration and return to safe independent feeding markedly exceeds the improvements typically seen in the chronic stage of TBI. Similar considerations apply to facial paresis and speech.^4,42–44^ Indeed, TBI01 exhibited substantial improvements in vocal intensity, and phonation after 4 years since initial injury, 3 years after DBS implantation, far exceeding the typical recovery trajectory of post-TBI dysarthria. Finally, DBS also improved cervical posture, reversing FHP and pathologic CVA that has been shown to negatively impact swallowing biomechanics and vocal phonation.^31,45–47^ This restoration of TBI01’s cervical posture, atypical to natural recovery history reported for severe TBI, mediated in part by the continuous motor thalamus stimulation effect on neck and paraspinal muscles spasticity may have supported functional improvements observed across swallowing and speech.

Importantly, the long-term effects of stimulation differed across motor functions. Facial symmetry, swallowing, vocal intensity, phonation, and speech efficiency improved progressively over time and persisted in the absence of stimulation. In contrast, articulation with DBS OFF fluctuated with speech therapy status, and optimal articulatory performance was maintained only with DBS ON or during periods of concurrent speech therapy. Moreover, while performance with DBS ON continued to improve over time for gross motor functions, articulatory performance with DBS ON remained stable across years, suggesting that chronic stimulation did not raise the ceiling of fine articulatory control beyond its immediate assistive effect. One possible explanation is that functions such as swallowing and phonation rely on bilateral cortical representation and brainstem neural activity,^48^ which may provide sufficient residual substrate for long-term reinforcement. In contrast, corticobulbar projections to the major muscle of tongue and lips, are predominantly contralateral, making them more vulnerable to lesions. Moreover, fine articulation requires fast, precise, and individuated control of multiple articulators. If so, the extent of recovery of fine motor skills may be constrained by the amount of spared white matter, and combining stimulation with targeted behavioral therapy may be necessary to optimize these functions.

Our outcomes contribute to a growing body of literature investigating the potential of neuromodulation to promote functional recovery after cerebral injuries. Previous works have explored the use of non-invasive neuromodulation interventions, such as transcranial magnetic stimulation (TMS) and transcranial direct current stimulation (tDCS),^49–52^ to increase cortical excitability and improve dysarthria and dysphagia, yet results have been inconsistent and rarely extend beyond one year post-injury.^17,53,54^ Instead, our approach showed that continuous DBS promotes long-lasting improvements across orofacial motor control, swallowing, and speech up to 4 years post-injury. These more robust effects may reflect a greater anatomical selectivity and continuous nature of DBS relative to non-invasive approaches. Our results also support prior DBS investigations reporting improvement in cognitive and upper-limb motor functions in patients with TBI.^55–57^ However, these studies did not examine the evaluation of acute assistive effects of stimulation and its capacity to promote long-term therapeutic gains, as we do here. Our findings therefore offer novel evidence for a dual role of motor thalamus DBS: as both an immediate facilitator of motor function and a long-term therapeutic driver of sustained recovery in patients with chronic orofacial motor deficits from TBI.

Despite these encouraging findings, the single-case design limits generalizability, and the broader applicability must be interperated cautiously until replicated in larger cohorts. The open-label nature of DBS also makes complete blinding of stimulation from the participant difficult. Since turning OFF the stimulation for extended periods of time would significantly reduce TBI01’s quality of life, we were unable to ethically conduct a prolonged DBS OFF control period to fully assess washout effects. This limitation raises the possibility that improvements observed during the brief DBS OFF settings reflect a lingering residual effect of stimulation rather than permanent clinically meaningful improvement. However, two observations argue against this interpretation. First, although the length of the OFF condition remained the same across years, the difference between stimulation ON and OFF was significantly larger in year 1 for most tests relative to later years, whereas a simple residual effect would be expected to produce a consistent difference over time. Second, a purely residual-effect explanation cannot account for the continued improvement in performance with stimulation ON, nor the corresponding reduction in the magnitude of the assistive DBS ON-versus-OFF effect that we observed. A related possibility is that the observed improvements reflect learning or practice effects from repeated testing over the three-year period, rather than a genuine therapeutic effect from stimulation. However, such practice-induced improvement would be expected to produce a consistent, monotonic improving trend across all testing years. This would not account for the decline in speech articulation performance we observed during year-2, which instead suggests that stimulation factors beyond simple repeated exposure to the testing paradigm may be driving the changes in performance over time. Nevertheless, larger cohort studies with proper blinding from stimulation parameters, appropriate control for conventional rehabilitation factors will be necessary to confirm these long-term therapeutic effects. Lastly, whether earlier implantation would yield greater benefit also remains unknown. TBI01 was implanted one year after injury, in the chronic phase, after the period of fastest spontaneous recovery. Stimulation during the acute or subacute phase, when the brain is actively adapting to injury, may produce larger, more durable gains. Reassuringly, the progressive gains observed here years after injury suggest that meaningful changes remains accessible in the chronic stage.

In conclusion, this study provides first-in-human evidence that continuous motor thalamus DBS can elicit long-term therapeutic effects, producing clinically significant recovery of facial volitional movement, swallowing, speech, and cervical posture in the chronic phase of TBI with DAI. The clinical implications of these findings extend beyond TBI to a broader spectrum of neurological disorders characterized by disruption of corticobulbar and corticospinal tracts. The motor thalamus occupies a strategic position within distributed motor circuits, integrating cerebellar and pallidal inputs and modulating cortical excitability through dense thalamocortical projections.^9,10^ As such, targeted low-frequency stimulation may represent a strategy to facilitate structurally preserved but functionally dormant pathways in conditions such as ischemic or hemorrhagic stroke, hypoxic–ischemic injury, tumor-related white matter disruption, and selected neurodegenerative disorders affecting bulbar function. Importantly, this approach reframes DBS not merely as a symptomatic therapy, but as a neurorestorative intervention potentially capable of promoting motor recovery well beyond traditional therapeutic windows.

## Methods

### Ethical Considerations

This study, which included functional tests in a human subject, was approved by the University of Pittsburgh Institutional Review Board (STUDY2007011) and Clinicaltrials.gov (NCT07056348). All the recordings were carried out according to CARE guidelines and in compliance with the Declaration of Helsinki principles. The participant was informed of the procedure and they signed an informed consent, which included the consent for the use of all data collected during the experiment in scientific publications. Additionally, participant provided written consent to the use of photos, videos, and voice-recorded acquired during the experiments for research purposes and publications.

### Human participants

This is a case study where we followed one subject, TBI01, across the span of 3 years (see **Supplementary Table 1** for clinical details and experiments performed). TBI01 sustained a severe traumatic brain injury resulting in DAI from a motor vehicle accident around 1 year before DBS implantation. DBS was implanted in the interface of the ventralis intermediate and ventralis oralis posterior (VIM/VOP) nuclei of the motor thalamus to treat medically-intractable asymmetric post-traumatic action tremor symptoms that developed after the injury **(Figure 1B).** The DAI involved the CST and CBT and was accompanied by bilateral edema of the cerebral peduncles and pons (**Figure 1A**). Consequently, TBI01 sustained paresis of the right side and left upper extremity muscle weakness and tremor with decreased overall strength, balance, coordination, and endurance. According to SLP evaluations, the participant suffered profound motor speech impairment characterized by slow, uncoordinated, and imprecise speech with overall poor comprehensibility in unknown listeners’ context. The SLP also noted probable dysphonia with reduced vocal intensity, and hoarse vocal quality likely secondary to reported left vocal fold paralysis. With regard to swallowing function, the participant demonstrated moderate oropharyngeal dysphagia characterized by suspected oral phase discoordination and possible airway compromise due to left vocal fold paralysis. The participant was recommended for bilateral DBS electrode implant (Medtronic, Sensight leads Percept PC) in VIM/VOP nucleus to treat the post-traumatic tremor. Stimulation tests were conducted stimulating all contacts located within the VIM/VOP (i.e., without current steering).

### Diffusion MRI and tractography analysis

Diffusion images for TBI01 were acquired on a SIEMENS Prisma Fit scanner using a diffusion sequence (2mm isotropic resolution, TE/TR= 99.2 ms/2490 ms, 257 diffusion sampling with maximum b-value 4010 s/mm²). The accuracy of b-table orientation was examined by comparing fiber orientations with those of a population-averaged template. ^58^ The tensor metrics were calculated using diffusion weighted images (DWI) with b-value lower than 1750 s/mm².

To quantify the white-matter fibers damage in TBI01, we used an advance diffusion MRI technique commonly used in patients with white-matter damage that allows for quantification of a decrease in anisotropic diffusion by applying a “tracking-the-difference” paradigm when compared to a normal population template: differential tractography. ^59^ Differential tractography was performed by comparing the high-definition fiber tractography (HDFT) of TBI01 against an age-and-sex-matched template constructed from a group of healthy controls of healthy fractional anisotropy (FA). To capture neural injury, we highlighted tracks that displayed a 10% decrease in FA when compared to the template of healthy controls. We then quantified the composition of lesioned tracts based on a population-averaged atlas of human white matter tracts for the main fiber tracts: corticospinal tract, corticobulbar tract, and dentato-rubro-thalamic tract.^58^

### Anatomical localizaiton of DBS electrodes

Implanted DBS electrodes were localized using Lead-DBS (v3.2.1). ^60^ Postoperative CT image was linearly coregistered to preoperative T1-weighted MRI using Advanced Normalization Tools (ANTs).^61^ To account for brain shift caused by pneumocephalus and electrode insertion, coregistration was refined with the brainshift correction module, using a coarse subcortical mask.^62^ The preoperative image was then nonlinearly normalized to MNI152NLin2009bAsym template space (ICBM 2009b nonlinear asymmetric)^63^ using the symmetric diffeomorphic (SyN) registration approach implemented in ANTs,^61^ with the preset parameters. Where needed, minimal manual corrections of the thalamic borders were applied with the WarpDrive tool, ^64^ using the whole-thalamus segmentation of the THOMAS atlas as the anatomical reference. ^65^ Individual thalamic nuclei were subsequently defined using the DISTAL atlas. ^66^ Electrode trajectories and contacts were automatically identified on postoperative CT using the PaCER algorithm.^67^ Because directional leads were implanted, lead rotation was set manually in Lead-DBS according to the orientation estimates provided by the manufacturer’s reconstruction. The volume of tissue activated (VTA) was estimated using the finite element method implemented in the SimBio/FieldTrip pipeline, ^68^ with default parameter for conductivities and threshold and using DISTAL atlas for grey matter identification. Finally, all electrode reconstructions, VTAs, and atlas structures were rendered in MNI space for visualization.

### Behavioral tests

TBI01 performed a series of functional tests to assess and track progress on impairments over multiple sessions whenever possible (see **Supplementary Table 2**). All functional tests were repeated without and with motor thalamus stimulation. For each session, the amplitude and frequencies of motor thalamus stimulation were selected to be at 55Hz which would produce a transient paresthesia in the facial muscles (see **Supplementary Table 1** for details on stimulation parameters). Tests were initiated only when TBI01 reported that the paresthesia disappeared. For all tasks, a computer monitor was placed in front of the participants, about 25-30 inches away from their faces to instruct them on the tasks. TBI01 was instructed to initiate the trial after a cue that paired a short tonal sound with the appearance of the action prompt on the screen. For tasks with time constraints, participants received feedback on the remaining time during the trial and visual feedback to stop the instructed action after the allotted time. A directional microphone was placed in front of the participant to record both the participants’ speech and the auditory go cue from the monitor without occluding the behavioral task displayed on the monitor. TBI01 completed all behavioral tasks sitting in a chair. Stimulation ON and OFF conditions were randomized across sessions.

#### Facial tasks

To evaluate the stimulation effect on voluntary facial movement, four facial tasks were designed to evaluate the range of movement and aperture of facial articulators critical to speech and swallowing: 1) protruding tongue in/out (TT); 2) smiling (TS); 3) opening/closing mouth (TO); 4) puckering the lips (TL). For all tasks, TBI01 was instructed to alternate between a neutral facial expression and the designated facial task as fast and as large as they could within 15 seconds. No visual feedback was provided to prevent the subject from adjusting their movements between conditions. Video of the facial movements were recorded. Only sessions where TBI01 could complete the tasks for all stimulation conditions while sitting in a consistent upright posture were included (see **Supplementary Table 2**).

#### Single words

We performed tests to measure the effects of stimulation on articulation. TBI01 was tasked to repeat single words five times as clearly as possible. We tested words composed of one to three syllables (n = 37 one syllable, n = 20 two syllables, and n = 2 three syllables) and ensured that all American English phonemes were tested in at least one experimental session. TBI01 performed this task across multiple sessions. Only sessions in which TBI01 could complete all tasks for all stimulation conditions were included in the data analysis (see **Supplementary Table 2**).

#### Sentences

We performed tests to measure the effects of stimulation on speech phonation, intelligibility, and quality. TBI01 was tasked to repeat three times aloud short (5 to 8-word) phoneme-balanced sentences selected from the Assessment of Intelligibility of Dysarthric Speech test (AIDS). TBI01 performed these tasks across multiple sessions in year 1 and year 3, while audio recordings were acquired. Only sessions in which the participant could complete all tasks for all stimulation conditions were included in the analysis (**Supplementary Table 4**).

#### Perceptual Speech Assessment

Speech quality was assessed on the sentences. All audio recordings were processed via Adobe Premiere Pro and MATLAB to blind across sessions. We then employed a validated auditory-perceptual assessment of dysarthria through Qualtrics. Audio files of sentence samples from TBI01 were provided to n=6 naive native English-speakers blinded to session temporality. Evaluators were instructed to rate the following perceptual metrics on a sliding scale of 0 to 100, from most to least impairment, respectively. To ensure intra-rater reliability, rater response was included only if repeated ratings within each metric differed by <30%.^69,70^ Grades were then compared between year 1 and year 3 for the following perceptual metrics:

1. Speech Intelligibility: Judge how well the speech is understood.
2. Listener Effort^35,36^: How effortful was it for you to understand this speaker?
3. Speech Naturalness^37^: Rate the naturalness of speech

a. Pay attention to how the speech compares with that of non-disordered speech as well as if it conforms to standards of rate, rhythm, intonation, and stress patterning and if it conforms to the grammatical structure of the utterance being produced.
4. Articulatory Precision: Judge how precise the speech is; scale severity based on whether some sounds vs. most sounds were slurred.
5. Speech Rate: Rate severity based on whether speakers are somewhat slow vs. very slow.
6. Overall Voice Quality: Rate severity based on how the voice compares with that of non-disordered voice (e.g., breathiness, roughness, harshness, etc.)
7. Overall Speech Severity^42^: Rate the overall severity of speech paying attention to the following:

a. Voice (quality – breathy, noisy, gurgly, high pitch, too low pitch, or sounds normal)
b. Resonance (too nasal, not nasal in the right places, sounds like they have a cold, or sound normal)
c. Articulatory precision (some sounds are crisp or slurred or somewhere in between or sounds normal)
d. Speech rhythm (the timing of speech doesn’t sound right or sounds normal)
e. Pay attention to overall speech naturalness and prosody (melody and timing of speech). Do not focus on the speaker’s intelligibility or how understandable each sentence is.

#### Videofluoroscopic swallow study^13^

Prior to phase-2 of DBS implant (1-year post-TBI), TBI01 completed a videofluoroscopic swallow study to assess the severity of swallowing impairment. At the time of this initial assessment, TBI01 was on a soft and bite-sized diet with mildly thick liquids (IDDSI level 6 and level 2, respectively). Adhering to these reports, the videofluoroscopic swallow study (i.e., modified barium swallow, MBS) consisted of challenging the participant with the following adjusted consistencies: 1) mildly thick liquid via a straw, 5ml bolus; 2) unmeasured free straw sip; 3) pudding. By standard protocol, trials consisting of swallowing varying volumes of mildly thick liquid (5ml and unmeasured free straw sip) were repeated twice. Importantly, the participant was instructed to clear the bolus in a single swallow. Therefore, any portion of the bolus remaining after the first swallow was considered as residue. Videofluoroscopy was acquired for each swallow at a frequency of 60 frames per second. Due to coordination limitations and the radiation exposure, the same test was repeated only after DBS implant (3 years post-TBI) with and without motor thalamus stimulation at 55 Hz to assess the progress overtime. Due to improvement in diet status by year- 3, TBI01 successfully completed all trials per DIGEST protocol; however, only trials consistently performed at both year 1 and 3 were included for matched-comparison analysis. SLPs (n=2) blinded from the stimulation conditions was then tasked with scoring all swallow trials of the study based on the Penetration Aspiration Scale (PAS) ^14^ and the Digesting Image Grade of Swallowing Toxicity (DIGEST) score.^15^ Importantly, since DIGEST scoring is by definition only evaluated from the first swallow, spillage of bolus remaining after the initial swallow was considered “residue” even if remnants were cleared by consequent swallows. The quadratic weighted Cohen’s kappa was calculated to assess interrater reliability between the 2 SLPs scoring for both the PAS and DIGEST. We constructed a confusion matrix consisting of both SLPs ratings from which we computed the quadratic weights to create an observed proportion matrix representative of interrater discrepancies. An expected agreement matrix was then computed assuming both SLPs rated via random. We then computed the weighted discrepancies, kappa (k), between observed and expected matrices.^71,72^ k-value should be interpreted as follows: < 0.20 as poor agreement, 0.20–0.40 as fair agreement, 0.41–0.60 as moderate agreement, 0.61–0.80 as good agreement, and > 0.80 as very good agreement.

### Kinematic recordings and analysis

Kinematics of facial tasks were recorded by a GoPro® Camera and analyzed through DeepLabCut, a neural network for 2D and 3D markerless pose estimation based on transfer learning.^73^ All videos recorded of TBI01 performing facial tasks were initially processed using Adobe Premiere Pro to standardize framing and lighting across stimulation conditions. Kinematic facial articulators were manually labeled for a minimum of 40 frames per video clip. The labeled facial structures included jaw, cheek, mouth, upper lip, lower lip, nose, chin, and chest. We employed DeepLabCut and a cloud computing cluster (Google Colab Pro) to train a neural network and extrapolate the articulator positions across all frames. Each dataset for each participant was trained for a minimum of 300,000 iterations. Visual inspection from LT confirmed facial articulators automatically labeled by trained DeepLabCut models. Final x- and y-coordinates of each articulator across all tasks were used for kinematic analysis. For each movement repetition, movement initiation and completion were identified manually from visual inspection of the kinematic traces and videos.

#### Facial Tasks

We quantified mouth aperture of the left and right face at maximal facial task activation by quantifying the area between three representative facial articulators (corner of mouth, upper lip, and lower lip) from each side, respectively using ImageJ. Apeature values across time were scaled to intercanthal distance of 35mm. We then calculated the ratio between the right relative to the left mouth apertures as a measure of facial asymmetry (ratio = 1, no asymmetry; ratio < 1, asymmetry).

#### Posture

We calculated the chin-to-chest distance from the average of 20 sets of x- and y-coordinators of the chin and chest articulators randomly sampled for each testing session throughout the facial tasks test. The mean chin-to-chest distance was calculated across sessions by year, scaled to interchanthal distance of 35cm.

### Analysis of videofluoroscopic swallow study

In addition to the SLP evaluation of the videos, we performed a quantitative analysis of the swallow duration and maximum amount of residue. Specifically, swallow duration was calculated by the number of frames in the radiology video report from the first observation of modified barium within the oral cavity to the end of the swallow, defined by complete clearance of all contents uptaked. Only trials where consistent phases (oral, pharyngeal, or esophageal) were captured in the radiology report for both year 1 and year 3 were included. Behavioral signs indicative of swallowing completion included clearance of barium through the upper esophageal sphincter and relaxation of the sphincter to its resting state. The Area of Residue was quantified through ImageJ. Because the instructions provided to the subject were to clear the bolus in a single swallow, the area of residue was calculated after the first swallow. Specifically, one representative frame capture for each phase - matched trial with the maximal visible modified barium bolus was manually labeled as Region of Interest (ROI) from the videofluoroscopic swallow radiology video report. The total area of ROIs was calculated in Pixel^2^.

### Audio Recordings and analysis

All voice recordings were collected using a Sennheiser MKE 600 microphone (Sennheiser electronic SE & Co. KG, Germany) that was then amplified through a M-track Duo (48 kHz, 2-channel USB audio interface with 2 Combo Inputs, Crystal Preamps, and Phantom Power, M- audio, Rhode Island, USA) and finally digitally processed, and recorded using the Ripple Neuro Grapevine and Trellis software at a sampling frequency of 30 kHz. Each audio file was then cut into single words using a semi-automatic MATLAB script that removed pauses and non-task relevant sounds. The cutting process was verified by both auditory and visual inspection before the cut words were saved as both MATLAB structures and WAV files. To determine the beginning and end of individual phonemes, we used the Montreal Forced Aligner (MFA) to train an acoustic model to align phoneme transcripts to audio files. Specifically, we used the CMU ARPABET library ^74^ to train the model as it is a dictionary for North American English. Models were trained specifically to TBI01 including all sessions. The final output of the model was grapheme “TextGrids” compatible with PRAAT ^75^ Phoneme alignment was then inspected and confirmed for each WAV file using a custom PRAAT script. The final grapheme information was added to the MATLAB structures, storing both single words and their individual phoneme traces. All MATLAB analysis used these structures and all PRAAT analysis used the WAV and TextGrid datasets.

### Speech Assessment

Speech analysis was performed across the single words and sentences tasks to measure stimulation effect on speech subsystems and metrics. Due to the profound deficits of TBI01, we always excluded the first trial of each single word. This was to reduce any confounding/misleading effects from initiation variability and errors, including coughing, speaking over the tonal go cue, poor breath support, and whispering the word. Each word within a session was considered as a single word, resulting in a dataset of 108 words. We calculated several speech metrics to assess the effects of the stimulation on speech production.

#### Speech Intensity

To assess changes in speech intensity with motor thalamus stimulation, we first decomposed the audio signal of every recorded word into a spectrogram. The spectrogram was calculated every 5 ms using a 25 ms Hanning window and the frequency range was limited to 50-10000 Hz. We calculated the total power in decibels for each time step and then applied a second-order 1D median filter to the trial trace. For each trial, we calculated speech intensity as the median value of the smoothed trace. A standardized tone of consistent volume of 75 dB was used to calibrate microphone gains and acquisition settings at the beginning of each session.

#### Voice Breaks

To evaluate the effects of the stimulation on phonation, we calculated the number of voice breaks detected within each sentence using PRAAT’s voice report function. We first calculated a pitch object using a filtered autocorrelation method (recommended for voice pathology) over the entire sound bit with the standard settings save for pitch range which we adjusted to 75-400 Hz and the voice threshold which we reduced to 0.25 to better evaluate irregularities in the speech signal. We then created a PointProcess object to define the pulse train for the signal before running PRAAT’s voice report function with its standard settings. Voice breaks were defined as any inter-pulse intervals between voiced frames that lasted longer than 16.67ms.^10^

#### Consonant Articulation

To determine whether there was improvement in the articulation of consonants, we perceptually assessed the presence and quality of each consonant. For TBI01, two researchers (EG and LT) blinded to both the stimulation condition and session listened to the word while being presented with a transcript of the word and its phonemes. The researcher then classified each phoneme as being 1) *present*, 2) *distorted,* or 3) *fully absent*. Phoneme distortions were classified as pronunciation of incorrect phonemes (e.g. /tæk/ instead of /tɪk/). Next, we calculated the percentage of present and distorted/absent phonemes for stimulation OFF and stimulation ON for each session by concatenating the phonemes across words. To account for the confounding effect of fatigue, results from sessions with matched testing condition order (stimulation ON first, then stimulation OFF) were selected and plotted over time. Notably, 3 testing sessions conducted during year 2 were conducted virtually, thus were only exclusively included for consonant articulation analysis but excluded from vocal intensity analysis.

#### Speech Rate

To assess whether stimulation improves speech rate, we recorded the total duration of time required for TBI01 to complete single words speech tasks across DBS OFF and ON conditions normalized to the number of words repeated across all sessions to carculate utterance time in seconds per word. The less time required to complete testing suggest faster speech rate.

### Craniovertebral Angle Analysis

To calculate the craniovertebral angle (CVA) change over time, we acquired x-ray imaging for TBI01’s cervical spine at year 1 and year 3 in a supine position, with the back of the head in contact with the imaging table as much as possible. The Images were processed through ImageJ to quantify the angle between a horizontal line through the C7 vertebral level and a line from the C7 spinous process to the tragus of the ear.

### Endurance

We timed the duration spent in completing all testing within a session to assess stimulation’s effect on overall endurance. In all sessions, TBI01 was pushed to complete testing (facial tasks, speech, upper motor tasks) across various stimulation conditions until his maximum tolerance was reached upon his request to stop. Therefore, an increase in tolerated time suggested improvement in endurance on overall motor performance.

### Patient-Reported Outcomes

#### Qualitative Surveys

TBI01 were asked to complete surveys Global Rating of Change (GROC) and EQ-5D- 5L surveys annually for three years post-TBI. Percentage of improvement across time was calculated from initial scores.

### Statistical Procedures

All statistical comparisons of means presented in this manuscript were performed using the bootstrap method, a non-parametric approach which makes no distributional assumptions on the observed data. Instead, bootstrapping uses resampling to construct empirical confidence intervals for quantities of interest. For each comparison, we constructed bootstrap samples by drawing a sample with replacement from observed measurements, while preserving the number of measurements in each condition. We construct 10,000 bootstrap samples and, for each, calculate the difference in means of the resampled data. We employed two-tailed bootstrapping with alphas of 0.05 (95% confidence interval), 0.01 (99% confidence interval), or 0.001 (99.9% confidence interval). The null hypothesis of no difference in the mean was rejected if 0 was not included in the confidence interval of the corresponding alpha value. If more than one comparison was being performed at once, we used a Bonferroni correction by dividing the alpha value by the number of pairwise comparisons being performed. Nonparametric paired t-tests were used specifically for the analysis of speech fatigue. Fatigue analysis evaluates the time duration spent repeating the first and last five words tested within each session. Since the length of the words tested for each session was distinct, the time necessary to complete repetitions of the first and last five words varies. Hence, to evaluate the stimulation effect on speech fatigue, duration was paired within each session where the words tested were consistent between stimulation ON/OFF conditions.

## Data Availability

All data produced in the present study are available upon reasonable request to the authors

