## Supplementary materials for "Low-Frequency Deep Brain Stimulation Promotes Long-Term Orofacial Motor Recovery After Traumatic Brain Injury: A Case Study"

[+]These authors contributed equally

### Supplementary Materials:

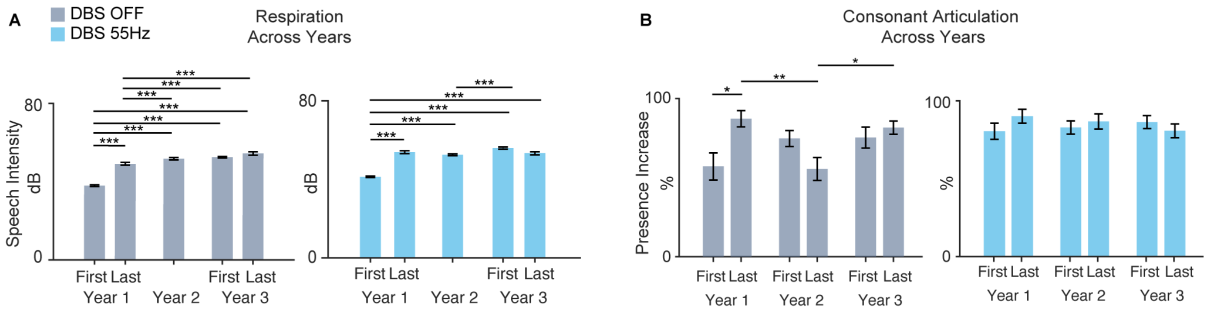


**Supplementary Figure 1 | Therapeutic Effects of DBS on Respiration and Articulation Across Years. (a)** Bar plot illustrating average speech intensity between first and last sessions for each year across three years post-DBS implant for DBS OFF and ON. Note, only 1 session was conducted in Year 2 as indicated. **(b)** Bar plot illustrating percentage of perceived consonants between first and last sessions of each year across three years post-DBS implant for DBS OFF and ON. For all panels, DBS OFF is labeled in gray and ON in blue. Statistical significance was assessed with two-tail bootstrapping with Bonferroni correction with $p<0.05$ (*), $p<0.01$ (**), $p<0.001$(***).

**Supplementary Table 1** - Stimulation Parameters

| | Motor Thalamus Stimulation Parameters | | |
|---|---|---|---|
| **Stimulation** | **Frequency (Hz)** | **Pulse Width** | **Amplitude (mA)** |
| Medtronic Bilateral VIM/VOP-DBS | 55 | 90us continuous | 3 |
| | 130 | 90us continuous | 1.5 |

**Supplementary Table 2** – Testing Sessions Timeline: Vocal Intensity session split from Phoneme

| | |
|---|---|
| **D-(-354)** | **Initial Injury** |
| D-(-41) | DBS Implant Stage I |
| D-(-34) | DBS Implant Stage II |
| **D-0** | **DBS Stim ON @ 55Hz** |
| Year 1 - D-126<br>Year 2 - D-415<br>Year 3 - D-761 | Facial expression task |
| Year 1 - D-(-39)<br>Year 2 - D-761 | Videofluoroscopic Swallow Study |
| Year 1<br>- D-106<br>- D-107<br>- D-123<br>- D-126<br>- D-131<br>- D-135<br>Year 2<br>- D-415<br>- D-513*<br>- D-616*<br>- D-644*<br>Year 3<br>- D-761<br>- D-937 | Single words speech task – phoneme & intensity analysis<br><br>Phoneme-balanced sentences – voice breaks & qualitative metrics<br><br>*phoneme analysis ONLY |
| Year 1<br>- D-107<br>- D-126<br>- D-135<br>Year 3<br>- D-897 | Phonation analysis – sentence tasks |
| Year 1 - D-186<br>Year 3 - D-938 | Cervical Radio-imaging |

**Supplementary Table 3 - MBS Scoring report** including PAS Scores, residue quantification, and DIGEST score of all swallowing trials evaluated by SLP (n=2) who are blinded to stimulation condition.

| | **Penetration Aspiration Scale** | | | **Pharyngeal Residue** | | | **DIGEST** | | |
|---|---|---|---|---|---|---|---|---|---|
| Trials | Year 1 Stim **OFF** | Year 3 Stim **OFF** | Year 3 Stim **ON** | Year 1 Stim **OFF** | Year 3 Stim **OFF** | Year 3 Stim **ON** | Year 1 Stim **OFF** | Year 3 Stim **OFF** | Year 3 Stim **ON** |
| **5mL**<br><br>IDDSI Lvl 4 | SLP1: 5<br><br>SLP2: 5 | SLP1: 1<br><br>SLP2: 1 | SLP1: 1<br><br>SLP2: 1 | SLP1: <10%<br><br>SLP2: <10% | SLP1: <10%<br><br>SLP2: <10% | SLP1: <10%<br><br>SLP2: <10% | SLP1: 1<br><br>SLP2: 1 | SLP1: 0<br><br>SLP2: 0 | SLP1: 0<br><br>SLP2: 0 |
| **5mL**<br><br>IDDSI Lvl 2 | SLP1: 1<br><br>SLP2: 1 | SLP1: 1<br><br>SLP2: 1 | SLP1: 1<br><br>SLP2: 1 | SLP1: <10%<br><br>SLP2: <10% | SLP1: 10%<br><br>SLP2: <10% | SLP1: <10%<br><br>SLP2: <10% | SLP1: 0<br><br>SLP2: 0 | SLP1: 0<br><br>SLP2: 0 | SLP1: 0<br><br>SLP2: 0 |
| **5mL**<br><br>IDDSI Lvl 2 | SLP1: 1<br><br>SLP2: 2 | SLP1: 1<br><br>SLP2: 1 | SLP1: 1<br><br>SLP2: 1 | SLP1: <10%<br><br>SLP2: <10% | SLP1: 10-49%<br><br>SLP2: 10-49% | SLP1: <10%<br><br>SLP2: <10% | SLP1: 0<br><br>SLP2: 0 | SLP1: 1<br><br>SLP2: 1 | SLP1: 0<br><br>SLP2: 0 |
| **Free Sip 1**<br><br>IDDSI Lvl 2 | SLP1: 2<br><br>SLP2: 1 | SLP1: 1<br><br>SLP2: 2 | SLP1: 1<br><br>SLP2: 1 | SLP1: 10-49%<br><br>SLP2: <10% | SLP1: 10-49%<br><br>SLP2: <10% | SLP1: <10%<br><br>SLP2: <10% | SLP1: 1<br><br>SLP2: 0 | SLP1: 1<br><br>SLP2: 0 | SLP1: 0<br><br>SLP2: 0 |
| **Free Sip 2**<br><br>IDDSI Lvl 2 | SLP1: 3<br><br>SLP2: 4 | SLP1: 2<br><br>SLP2: 2 | SLP1: 1<br><br>SLP2: 1 | SLP1: <10%<br><br>SLP2: <10% | SLP1: 10-49%<br><br>SLP2: 10-49% | SLP1: <10%<br><br>SLP2: <10% | SLP1: 1<br><br>SLP2: 1 | SLP1: 1<br><br>SLP2: 1 | SLP1: 0<br><br>SLP2: 0 |

**Table 4** – **Qualitative Survey Report** Self-reported by TBI01 across 3 years post-DBS implantation on Global Rating of Change (GROC) and EQ-5D-5L. For GROC the higher the score, the more significant the positive improvement in change. For EQ-5D-5L, the lower the score, the lower the impact of injury on quality of life, suggesting an improvement.

| | **Year 1** | **Year 2** | **Year 3** |
|---|---|---|---|
| **Global Rating of Change (GROC)** | +1<br>'A tiny bit better' | +3<br>'Somewhat better' | +5<br>'Quite a bit better' |
| **EQ-5D-5L** | 23/25 | 22/25 | 15/25 |
| Mobility | 5/5 | 5/5 | 4/5 |
| Self-care | 5/5 | 5/5 | 3/5 |
| Usual Activities | 5/5 | 5/5 | 3/5 |
| Pain & Discomfort | 3/5 | 2/5 | 2/5 |
| Anxiety & Depression | 5/5 | 5/5 | 3/5 |
